# Comparison of temporal changes in left atrial and left ventricular strain after septal myectomy, alcohol septal ablation, and cardiac myosin inhibitor

**DOI:** 10.64898/2026.03.02.26347409

**Authors:** In-Chang Hwang, Minjung Bak, Jiesuck Park, Sang Yoon Kim, Joon Chul Jung, Hong-Mi Choi, Hyoung Woo Chang, Jae Hang Lee, Yeonyee E. Yoon, Hyung Gon Je, Jun Sung Kim, Sang Hon Park, Cheong Lim, Goo-Yeong Cho, In-ho Chae, Kay-Hyun Park

## Abstract

**Aims:** Cardiac myosin inhibitors (CMIs) have emerged as an alternative to septal reduction therapy (SRT) for obstructive hypertrophic cardiomyopathy (oHCM). However, comparative data on the time-trajectory of myocardial functional adaptation after septal myectomy (SM), alcohol septal ablation (ASA), and CMI are lacking. We compared temporal changes in echocardiographic parameters including LV global longitudinal strain (LVGLS) and LA reservoir strain (LASr) across these treatment strategies.

**Methods and Results:** In this single-center retrospective cohort, symptomatic oHCM patients treated with SM (n=22), ASA (n=11), or CMI (n=47) underwent serial echocardiography with deep-learning–based automated strain analysis. Primary outcomes were temporal changes in LVGLS and LASr. Mixed-effects models adjusted for baseline clinical and echocardiographic variables were used to assess time-trajectories for up to 24 months. Treatment success rates were 86.4% (SM), 72.7% (ASA), and 93.6% (CMI). LVOT gradients were similarly reduced across groups. LVEF showed a subtle early decline after CMI (adjusted P-for-interaction=0.019). LVGLS gradually improved after SM and ASA but remained unchanged with CMI. LASr significantly improved after SM, showed minimal change after ASA, and demonstrated late attenuation beyond 9–12 months in the CMI group (adjusted P=0.029).

**Conclusions:** Despite comparable LVOT gradient reduction, myocardial functional adaptation differed across therapies. Conventional SRT was associated with progressive improvement in LV and LA strain, whereas CMI therapy showed stable LVGLS with subtle early LVEF decline and late attenuation of LASr. These findings underscore the importance of longitudinal deformation imaging during CMI therapy and support reappraisal of SRT in selected patients requiring durable long-term management.

## Introduction

Obstructive hypertrophic cardiomyopathy (oHCM) is characterized by left ventricular outflow tract obstruction (LVOTO), resulting from myocardial hypertrophy, systolic anterior motion of the mitral valve, and hyperdynamic ventricular contraction. Because LVOTO represents a major determinant of symptoms, exercise intolerance, and adverse clinical outcomes, relief of LVOTO has therefore remained a central therapeutic goal.(1) Conventional treatment strategies recommend pharmacologic therapy, including beta-blockers or non-dihydropyridine calcium channel blockers, as first-line treatment, followed by septal reduction therapy (SRT) in patients with persistent symptoms despite optimal medical therapy. Septal myectomy (SM) and alcohol septal ablation (ASA) are established forms of SRT with comparable efficacy in relieving LVOTO and improving symptoms.(2) However, both procedures are invasive and carry procedural risks, including perioperative mortality and permanent pacemaker implantation, and have traditionally been reserved for patients with refractory symptoms.

The introduction of cardiac myosin inhibitors (CMIs) has substantially reshaped the therapeutic paradigm of oHCM. Contemporary guidelines now endorse CMIs as an alternative to SRT in selected patients.(1) Nonetheless, concerns persist regarding their long-term impact on myocardial function. Because CMIs directly reduce sarcomeric contractility, excessive suppression may result in unintended systolic dysfunction. Although recent studies have demonstrated improvement in left ventricular (LV) and left atrial (LA) function with CMI therapy, a decline in LV ejection fraction (LVEF) to <50% has been reported in approximately 10% of patients.(3–5) In contrast, the effects of SRT on myocardial function are heterogeneous. Both SM and ASA produce localized septal injury with early attenuation of regional longitudinal strain, followed by variable recovery.(6, 7) While some studies report improvement in global LV function after SRT, others show minimal change or deterioration in systolic indices.(8–10) Conversely, LA function and volume generally improve following effective LVOTO relief.(9, 11)

Despite expanding treatment options, no study has directly compared the temporal evolution of LV and LA deformation across all three contemporary strategies—SM, ASA, and CMI. This comparison is clinically relevant, as myocardial deformation indices such as LV global longitudinal strain (LVGLS) and LA reservoir strain (LASr) provide sensitive measures of cardiac function beyond conventional LVEF. Accordingly, we compared longitudinal changes in myocardial function among patients with oHCM treated with SM, ASA, or CMI using serial echocardiography with deep-learning–based automated strain analysis validated in oHCM.(12–14)

## Methods

### Study Population

This retrospective observational study included consecutive patients with oHCM who were evaluated and treated at Seoul National University Bundang Hospital (SNUBH). Eligible patients had clinically significant LVOTO and persistent symptoms (New York Heart Association [NYHA] functional class II–IV) despite guideline-directed medical therapy. Patients who underwent SM or ASA were identified from institutional procedural databases. After the introduction and reimbursement of the CMI mavacamten in South Korea in 2024, treatment strategies at our institution shifted, and newly treated symptomatic oHCM patients received mavacamten as the first-line therapy. Patients were excluded if echocardiographic image quality was insufficient for strain analysis or if follow-up echocardiographic data were unavailable.

The study protocol was approved by the Institutional Review Board of SNUBH (B-2004-604-409), and the requirement for written informed consent was waived due to the retrospective design and use of anonymized data.

### Treatment strategies and indications

Before the availability of CMI in South Korea, treatment decisions for symptomatic oHCM were made by a multidisciplinary heart team in accordance with contemporary guidelines.(1, 15) Selection of SM or ASA was based on septal anatomy, surgical risk, and patient preference. SM was generally preferred in patients with suitable septal morphology and acceptable surgical risk, whereas ASA was considered in patients with appropriate septal perforator anatomy who were suboptimal surgical candidates. Following the introduction of mavacamten, pharmacologic therapy with CMI was adopted as the first-line treatment strategy for symptomatic oHCM at our institution, consistent with updated guideline recommendations.(1, 15)

### Study outcomes

The primary outcomes were temporal changes in LVGLS and LASr following treatment with SM, ASA, or CMI. Serial echocardiographic examinations were analyzed to evaluate early and late changes in myocardial deformation over time. LVOT gradients and conventional echocardiographic parameters were assessed as supportive secondary measures.

Primary analyses were restricted to patients with successful treatment, defined as a maximum LVOT pressure gradient <50 mmHg within 6 months after SM, ASA, or CMI initiation. As a sensitivity analysis, temporal changes in echocardiographic parameters were additionally evaluated in the entire cohort, irrespective of treatment success.

### Echocardiography

Echocardiography was performed using commercially available ultrasound systems equipped with 2–2.5 MHz transducers, and measurements were obtained in accordance with European Association of Cardiovascular Imaging guidelines.(16) LV dimensions, septal and posterior wall thicknesses were measured, and LV mass was calculated using the Devereux formula. LV mass index (LVMI) was derived by normalizing LV mass to body surface area. LVEF were calculated by Simpson’s biplane method from apical 2- and 4-chamber views. LA volume index (LAVI) was obtained by dividing LA volume by body surface area.

### Automated measurement of LA and LV strain

LVGLS and LASr were quantified using a deep learning–based automated strain analysis platform (Sonix Health Workstation, Version 2.0; Ontact Health Co., Ltd., Korea), which performs automated view classification, endocardial border detection, segmentation, and deformation parameter extraction.(12, 17–20) The performance and validation of this system in patients with HCM, including those undergoing SRT or CMI therapy, have been reported previously.(12, 14) LVGLS was calculated as the mean peak longitudinal strain derived from apical 4-, 2-, and 3-chamber views, referenced to the onset of the QRS complex. LASr was measured from the apical 4-chamber view and defined as the first positive peak strain during ventricular systole, referenced to the QRS complex.

### Statistical Analysis

Continuous variables are presented as mean ± standard deviation or median (interquartile range), as appropriate. Categorical variables are expressed as counts and percentages. Temporal trajectories of echocardiographic parameters—including LVOT gradient, LVMI, LVEF, LAVI, LVGLS, and LASr—were modeled using generalized additive mixed models (GAMMs) with restricted cubic splines to account for nonlinear time effects and repeated measures within individuals. Models included treatment group, time, and their interaction as fixed effects, with patient identity specified as a random intercept. Multivariable adjustment included age, sex, baseline comorbidities (hypertension, diabetes mellitus, atrial fibrillation, coronary artery disease, chronic kidney disease, and prior stroke), and baseline echocardiographic parameters (LVOT gradient, LVEF, LVMI, and E/e′ ratio). For descriptive visualization and interval-based comparisons, echocardiographic parameters were summarized across predefined time intervals: baseline, <1 month, 1–3 months, 3–9 months, 9–15 months, and 15–24 months after SM, ASA, or CMI initiation. A two-sided P value <0.05 was considered statistically significant. All statistical analyses were performed using R version 4.3.2 (R Foundation for Statistical Computing, Vienna, Austria).

## Results

### Baseline characteristics

Baseline clinical and echocardiographic characteristics according to treatment strategy are summarized in **Table 1**. Patients in the SM group were younger than those in the ASA and CMI groups (60.6±13.4 vs. 68.3±12.2 vs. 64.8±14.6 years, respectively). The SM group more frequently had a family history of HCM and sudden cardiac death and demonstrated higher estimated glomerular filtration rates compared with the other groups. The ASA group was the oldest and had the highest burden of comorbidities, including hypertension, diabetes mellitus, atrial fibrillation, and chronic kidney disease. Symptom severity differed across groups. In the CMI group, 83% of patients were in NYHA functional class II, whereas NYHA class III symptoms were more common in the SM (63.6%) and ASA (72.7%) groups.

**Table 1.** Baseline characteristics.

|  | **Surgical**<br>**myectomy**<br>**(SM; n = 22)** | **Alcohol septal**<br>**ablation**<br>**(ASA; n = 11)** | **Cardiac myosin**<br>**inhibitor**<br>**(CMI; n = 47)** | **p-value** |
| --- | --- | --- | --- | --- |
| Age | 60.6±13.4 | 68.3±12.2 | 64.8±14.6 | 0.318 |
| Male sex | 10 (45.5%) | 3 (27.3%) | 29 (61.7%) | 0.089 |
| Body-mass index (kg/m<sup>2</sup>) | 27.6±5.3 | 24.7±2.8 | 25.9±3.6 | 0.115 |
| Systolic BP (mmHg) | 126.1±15.7 | 119.5±17.4 | 131.0±16.8 | 0.100 |
| Diastolic BP (mmHg) | 72.9±13.3 | 66.6±13.7 | 70.5±10.8 | 0.355 |
| Heart rate (bpm) | 64.8±16.1 | 66.3±11.8 | 68.2±10.5 | 0.566 |
| Chest pain | 6 (27.3%) | 6 (50.0%) | 2 (4.3%) | <0.001 |
| NYHA functional class |  |  |  | <0.001 |
| - II | 6 (27.3%) | 1 (9.1%) | 39 (83.0%) |  |
| - III | 14 (63.6%) | 8 (72.7%) | 7 (14.9%) |  |
| - IV | 2 (9.1%) | 2 (18.2%) | 1 (2.1%) |  |
| Hypertension | 11 (50.0%) | 10 (90.9%) | 28 (59.6%) | 0.070 |
| Diabetes mellitus | 9 (40.9%) | 6 (54.5%) | 16 (34.0%) | 0.441 |
| Atrial fibrillation | 3 (13.6%) | 2 (18.2%) | 5 (10.6%) | 0.779 |
| Coronary artery disease | 4 (18.2%) | 1 (9.1%) | 8 (17.0%) | 0.781 |
| Chronic kidney disease | 1 (4.5%) | 2 (18.2%) | 7 (14.9%) | 0.398 |
| COPD | 4 (18.2%) | 1 (9.1%) | 1 (2.1%) | 0.060 |
| Stroke | 0 (0.0%) | 1 (9.1%) | 4 (8.5%) | 0.363 |
| Family history of HCM | 9 (40.9%) | 2 (18.2%) | 9 (19.1%) | 0.129 |
| Family history of SCD | 6 (27.3%) | 1 (9.1%) | 1 (2.1%) | 0.005 |
| NSVT | 2 (9.1%) | 2 (18.2%) | 6 (12.8%) | 0.755 |
| Syncope | 5 (22.7%) | 2 (18.2%) | 2 (4.3%) | 0.057 |
| Prior ICD | 0 (0.0%) | 2 (18.2%) | 6 (12.8%) | 0.160 |
| LGE on CMR | 17/21 (81.0%) | 3/8 (37.5%) | 40/42 (95.2%) | <0.001 |
| Beta-blockers | 20 (90.9%) | 11 (100.0%) | 46 (97.9%) | 0.285 |
| NonDHP-CCB | 4 (18.2%) | 0 (0.0%) | 4 (8.5%) | 0.226 |
| Laboratory finding |  |  |  |  |
| NT-proBNP (pg/mL) | 1812.7 (906.5, 4294.2) | 2713.1 (2029.0, 3266.0) | 1045.4 (273.5, 2445.2) | 0.666 |
| Troponin I (ng/mL) | 0.04 (0.04, 0.11) | 0.03 (0.01, 0.16) | 0.05 (0.02, 0.14) | 0.896 |
| Serum creatinine (mg/dL) | 0.7 (0.6, 0.9) | 0.9 (0.8, 1.0) | 0.9 (0.8, 1.0) | 0.341 |
| GFR (mL/min/1.73m<sup>2</sup>) | 92.8 (81.2, 106.0) | 60.9 (58.2, 75.2) | 76.7 (63.3, 90.9) | 0.002 |
| Echocardiography |  |  |  |  |
| LVOT gradient (mmHg) |  |  |  |  |
| - at rest | 80.5 (38.4, 100.0) | 45.4 (23.0, 64.0) | 64.0 (25.0, 96.0) | 0.760 |
| - after provocation | 139.2 (92.2, 185.0) | 84.6 (72.3, 110.3) | 74.0 (57.8, 92.2) | 0.002 |
| LVEDD (mm) | 42.0 (40.0, 44.0) | 40.0 (38.0, 42.0) | 42.0 (38.5, 45.0) | 0.590 |
| LVESD (mm) | 23.0 (22.0, 26.0) | 24.3 (23.2, 26.6) | 25.0 (21.0, 27.5) | 0.899 |
| LVEF (%) | 65.3 (59.4, 71.2) | 65.9 (61.8, 68.0) | 68.4 (65.4, 70.8) | 0.176 |
| IVSd (mm) | 18.0 (16.0, 20.0) | 17.0 (16.0, 21.0) | 17.0 (14.5, 18.5) | 0.143 |
| LVPWd (mm) | 13.0 (11.0, 14.0) | 13.2 (11.0, 14.7) | 11.0 (10.0, 12.0) | <0.001 |
| LV max. thickness (mm) | 20.0 (19.0, 22.0) | 22.0 (19.5, 22.5) | 20.0 (17.0, 22.0) | 0.453 |
| LVMI (g/m<sup>2</sup>) | 145.1 (137.1, 194.4) | 146.8 (133.0, 166.9) | 126.0 (110.4, 140.5) | 0.003 |
| LAVI (mL/m<sup>2</sup>) | 69.8 (47.7, 82.1) | 58.4 (48.2, 79.4) | 48.7 (39.2, 63.4) | 0.012 |
| Mitral inflow E (m/s) | 0.80 (0.65, 0.91) | 1.05 (0.66, 1.17) | 0.66 (0.57, 0.84) | 0.044 |
| Mitral annular e' (cm/s) | 3.85 (3.30, 4.50) | 3.60 (2.61, 4.60) | 4.30 (3.35, 5.30) | 0.161 |
| E/e' ratio | 19.4 (18.7, 23.1) | 23.4 (13.8, 34.6) | 16.0 (12.2, 24.0) | 0.063 |
| TR Vmax (m/s) | 2.7 (2.3, 2.9) | 3.0 (2.6, 3.5) | 2.5 (2.3, 2.7) | 0.097 |
| PASP (mmHg) | 34.2 (26.2, 38.6) | 41.0 (32.2, 52.6) | 30.0 (26.2, 34.2) | 0.129 |
| LVGLS (%) | 13.6 (10.6, 15.8) | 15.9 (13.3, 17.0) | 14.2 (13.3, 17.0) | 0.140 |
| LASr (%) | 18.0 (14.0, 21.2) | 16.9 (12.8, 24.3) | 19.9 (17.1, 26.0) | 0.214 |

Values are shown in number (%) or median with interquartile ranges (Q1, Q3).
Abbreviations: BP, blood pressure; COPD, chronic obstructive pulmonary disease; HCM, hypertrophic cardiomyopathy; SCD, sudden cardiac death; NSVT, nonsustained ventricular tachycardia; ICD, implantable cardioverter–defibrillator; NonDHP-CCB, non–dihydropyridine calcium channel blocker; LGE, late gadolinium enhancement; CMR, cardiac magnetic resonance; NYHA, New York Heart Association; NT-proBNP, N-terminal pro–B-type natriuretic peptide; GFR, glomerular filtration rate; LVOT, left ventricular outflow tract; LVEDD, left ventricular end-diastolic dimension; LVESD, left ventricular end-systolic dimension; LVEF, left ventricular ejection fraction; IVSd, interventricular septal thickness at end-diastole; LVPWd, left ventricular posterior wall thickness at end-diastole; LVMI, left ventricular mass index; LAVI, left atrial volume index; TR, tricuspid regurgitation; PASP, pulmonary artery systolic pressure; LVGLS, left ventricular global longitudinal strain; LASr, left atrial reservoir strain.

The maximal provoked LVOT gradient was highest in the SM group, followed by the ASA and CMI groups (139.2 [92.2–185.0] vs. 84.6 [72.3–110.3] vs. 74.0 [57.8–92.2] mmHg, respectively; *p* = 0.002). LVEF was comparable among the three groups (65.3 [59.4–71.2] vs. 65.9 [61.8–68.0] vs. 68.4 [65.4–70.8]%, *p* = 0.176). LVMI was highest in the SM group, intermediate in the ASA group, and lowest in the CMI group (145.1 [137.1–194.4] vs. 146.8 [133.0–166.9] vs. 126.0 [110.4–140.5] g/m²; *p* = 0.003). Similarly, LAVI was greatest in the SM group and lowest in the CMI group (69.8 [47.7–82.1] vs. 58.4 [48.2–79.4] vs. 48.7 [39.2–63.4] mL/m²; *p* = 0.012). Markers of diastolic burden tended to be more pronounced in the ASA group, with higher mitral E velocity (1.05 [0.66–1.17] m/s, *p* = 0.044) and numerically higher E/e′ ratio (23.4 [13.8–34.6], *p* = 0.063) and TR Vmax (3.0 [2.6–3.5] m/s, *p* = 0.097). Baseline LVGLS did not differ significantly among groups (13.6 [10.6–15.8] vs. 15.9 [13.3–17.0] vs. 14.2 [13.3–17.0]%, *p* = 0.140). Likewise, baseline LASr was comparable (18.0 [14.0–21.2] vs. 16.9 [12.8–24.3] vs. 19.9 [17.1–26.0]%, *p* = 0.214).

### Treatment outcomes and reduction in LVOT gradient

Treatment outcomes are summarized in **Table 2** and **Figure 1**. During the 2-year follow-up, there were no cardiovascular deaths, sudden cardiac deaths, or ventricular arrhythmias. In the SM group, permanent pacemakers were implanted in three patients (13.6%), new-onset atrial fibrillation occurred in one patient (4.5%), and a new ventricular septal defect was detected in one patient (4.5%). No pacemaker implantation, new-onset atrial fibrillation, or ventricular septal defect occurred in the ASA or CMI groups. Among 22 patients undergoing SM, 19 (86.4%) achieved successful LVOT gradient reduction. Three patients had persistent severe LVOTO, two of whom subsequently received CMI therapy. In the ASA group, 8 of 11 patients (72.7%) achieved procedural success; two of the three patients with residual obstruction were later treated with CMI. In the CMI group, 44 of 47 patients (93.6%) achieved successful LVOT gradient reduction; one patient discontinued therapy due to chest discomfort, and two had persistent dynamic obstruction at the mid-LV or LVOT level.

**Figure 1.**
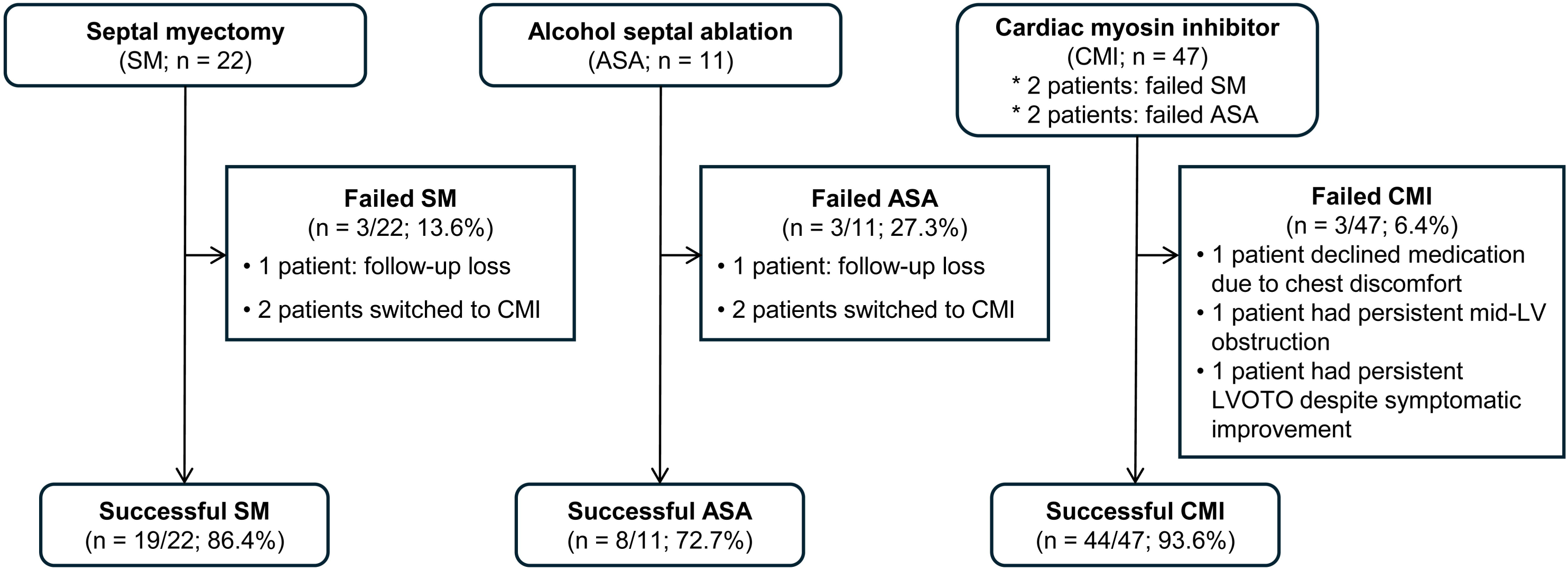
Flow chart of study population. Abbreviations: LV, left ventricular; LVOTO, left ventricular outflow tract obstruction.

**Table 2.** Treatment outcomes.

|  | **Surgical**<br>**myectomy**<br>**(SM; n = 22)** | **Alcohol septal**<br>**ablation**<br>**(ASA; n = 11)** | **Cardiac myosin**<br>**inhibitor**<br>**(CMI; n = 47)** |
| --- | --- | --- | --- |
| Cardiovascular death | 0 (0.0%) | 0 (0.0%) | 0 (0.0%) |
| Ventricular tachycardia/fibrillation | 0 (0.0%) | 0 (0.0%) | 0 (0.0%) |
| Pacemaker implantation | 3 (13.6%) | 0 (0.0%) | 0 (0.0%) |
| New-onset atrial fibrillation | 1 (4.5%) | 0 (0.0%) | 0 (0.0%) |
| New-onset ventricular septal defect | 1 (4.5%) | 0 (0.0%) | 0 (0.0%) |
| Treatment failure* | 3 (13.6%) | 3 (27.3%) | 3 (6.4%) |

\* Treatment failure was defined as the maximum LVOT pressure gradient <50 mmHg within 6 months after SM, ASA, or CMI.

Temporal analyses were restricted to patients with successful treatment. Both resting and provoked LVOT gradients significantly decreased after SM, ASA, and CMI (**Figure 2**). Although baseline gradients were highest in the SM group, resulting in a significant difference in overall trajectory (adjusted P-for-interaction = 0.005 by GAMM), post-treatment LVOT gradients were comparable among groups. In the CMI group, LVOT gradients exhibited mild fluctuations during follow-up, likely reflecting dose titration according to gradient severity and LVEF monitoring.

**Figure 2.**
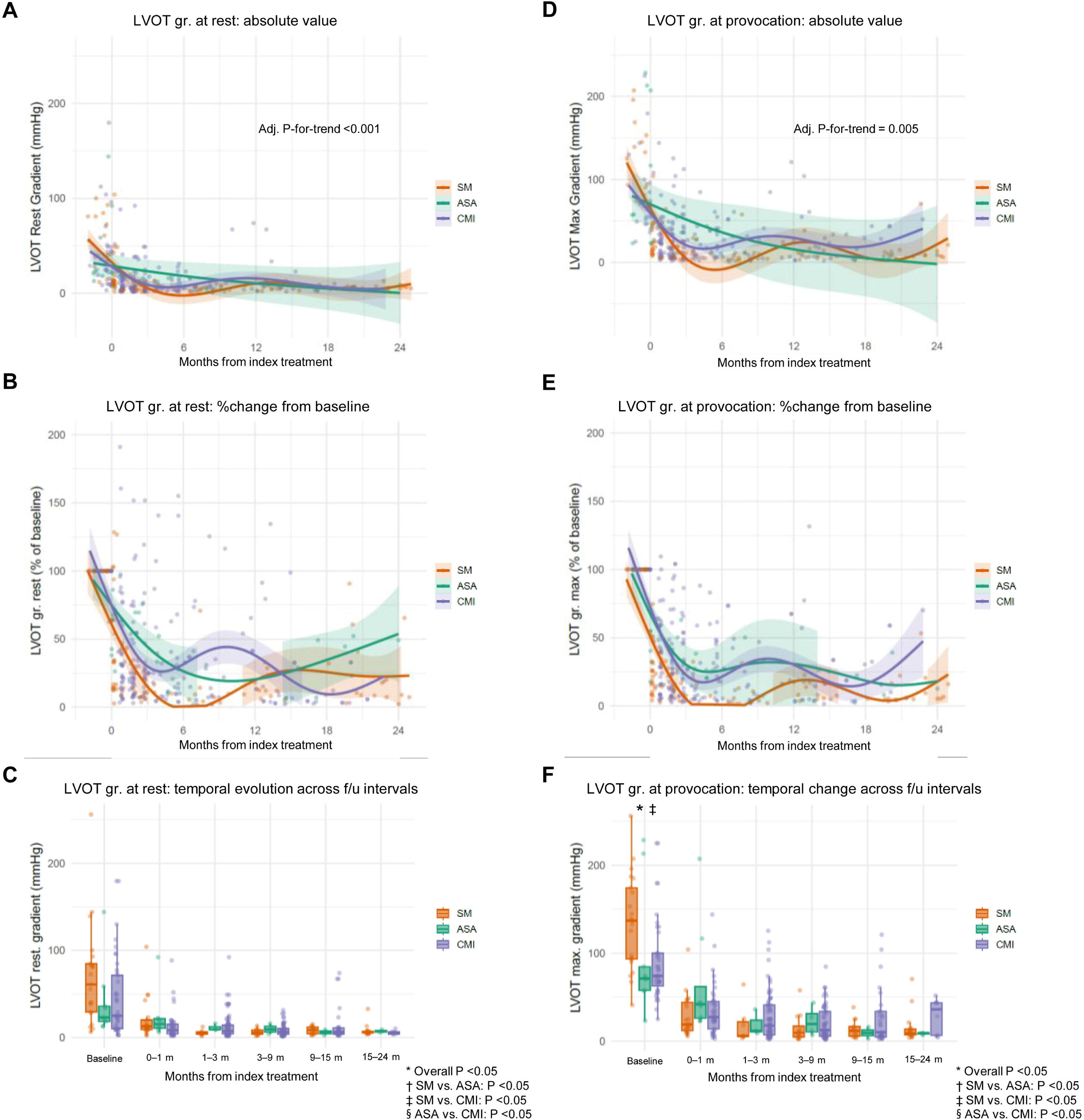
Time-trajectory of LVOT gradient. Temporal changes in LVOT gradient at rest assessed as its original values (A), in specific time intervals (B), and expressed as %change from baseline (C). Temporal changes in LVOT gradient after provocation as its original values (D), in specific time intervals (E), and expressed as %change from baseline (F). * Overall P values were calculated with generalized additive mixed models (GAMM), comparing the trajectory of echocardiographic variables with adjustment for age, sex, comorbidities at baseline (hypertension, diabetes, atrial fibrillation, coronary artery disease, chronic kidney disease, and stroke), and the baseline echocardiographic parameters (LVOT gradient, LVEF, LVMI, and E/e’ ratio).

### Temporal changes in LV function parameters

Temporal changes in LVEF and LVGLS are shown in **Figure 3**. When analyzed as absolute values, LVEF demonstrated a similar pattern of gradual and modest decline across groups (**Figure 3A**). However, a significant intergroup difference was observed in the adjusted trajectory (adjusted P-for-interaction = 0.019), driven by a subtle early decline in LVEF following CMI initiation when expressed as percentage change from baseline (**Figure 3B**).

**Figure 3.**
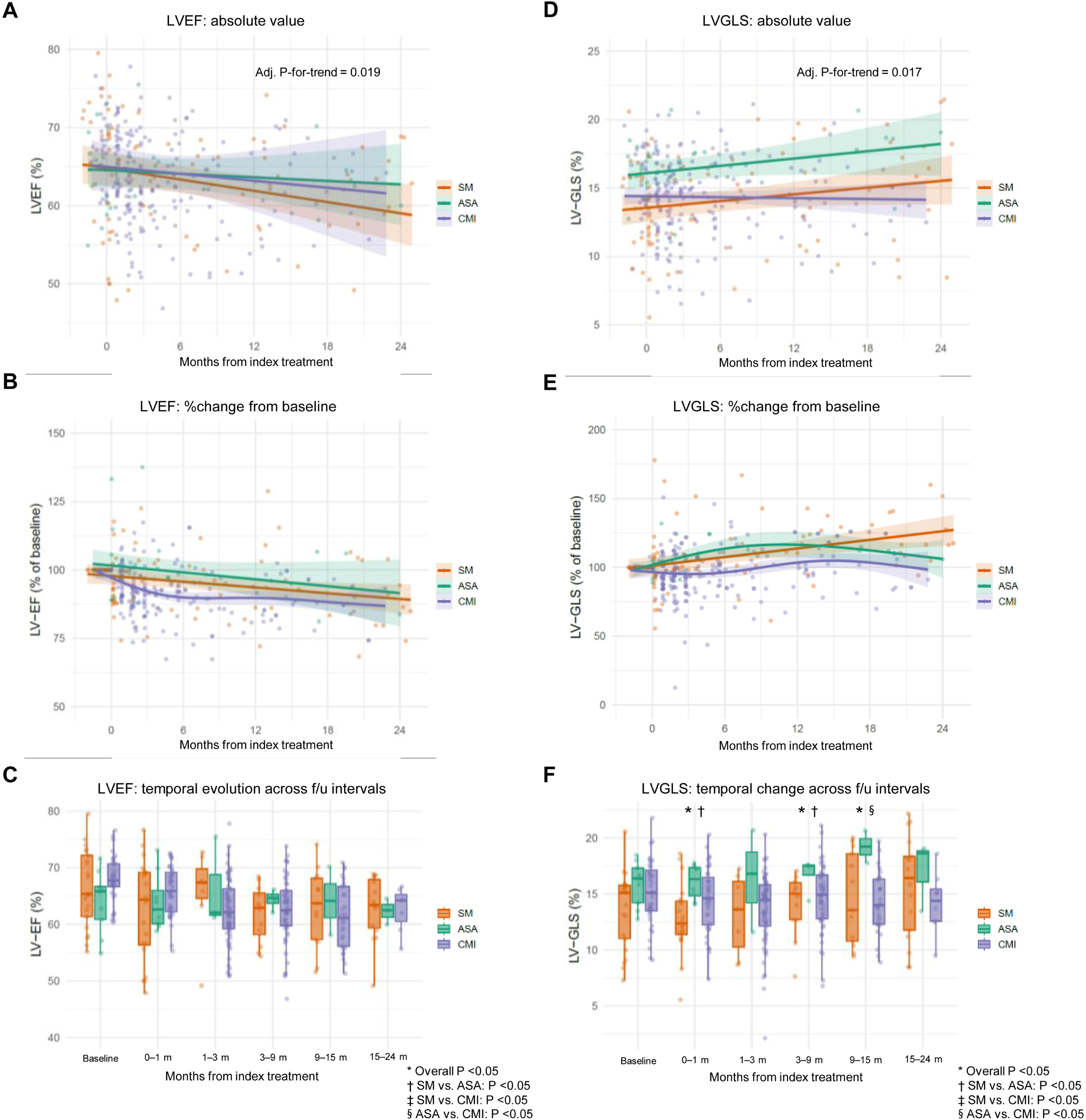
Time-trajectory of LV functional parameters. Temporal changes in LVEF assessed as its original values (A), in specific time intervals (B), and expressed as %change from baseline (C). Temporal changes in LVGLS as its original values (D), in specific time intervals (E), and expressed as %change from baseline (F). * Overall P values were calculated with generalized additive mixed models (GAMM), comparing the trajectory of echocardiographic variables with adjustment for age, sex, comorbidities at baseline, and the baseline echocardiographic parameters.

LVGLS improved progressively in the SM and ASA groups but remained largely unchanged in the CMI group (**Figures 3D–3E**). Interval-based analyses demonstrated significant intergroup differences during early follow-up, with higher LVGLS values observed after ASA compared with SM at <1 month and 3–9 months post-procedure. These differences were no longer present at 15–24 months.

### Temporal changes in LA volume and reservoir strain

The trajectory of LAVI differed when assessed using absolute values: LAVI gradually decreased in the SM and ASA groups but showed a mild increase in the CMI group (**Figures 4A–4C**). However, when expressed as percentage change from baseline and adjusted for baseline characteristics, intergroup differences were no longer statistically significant (adjusted P-for-interaction = 0.336).

**Figure 4.**
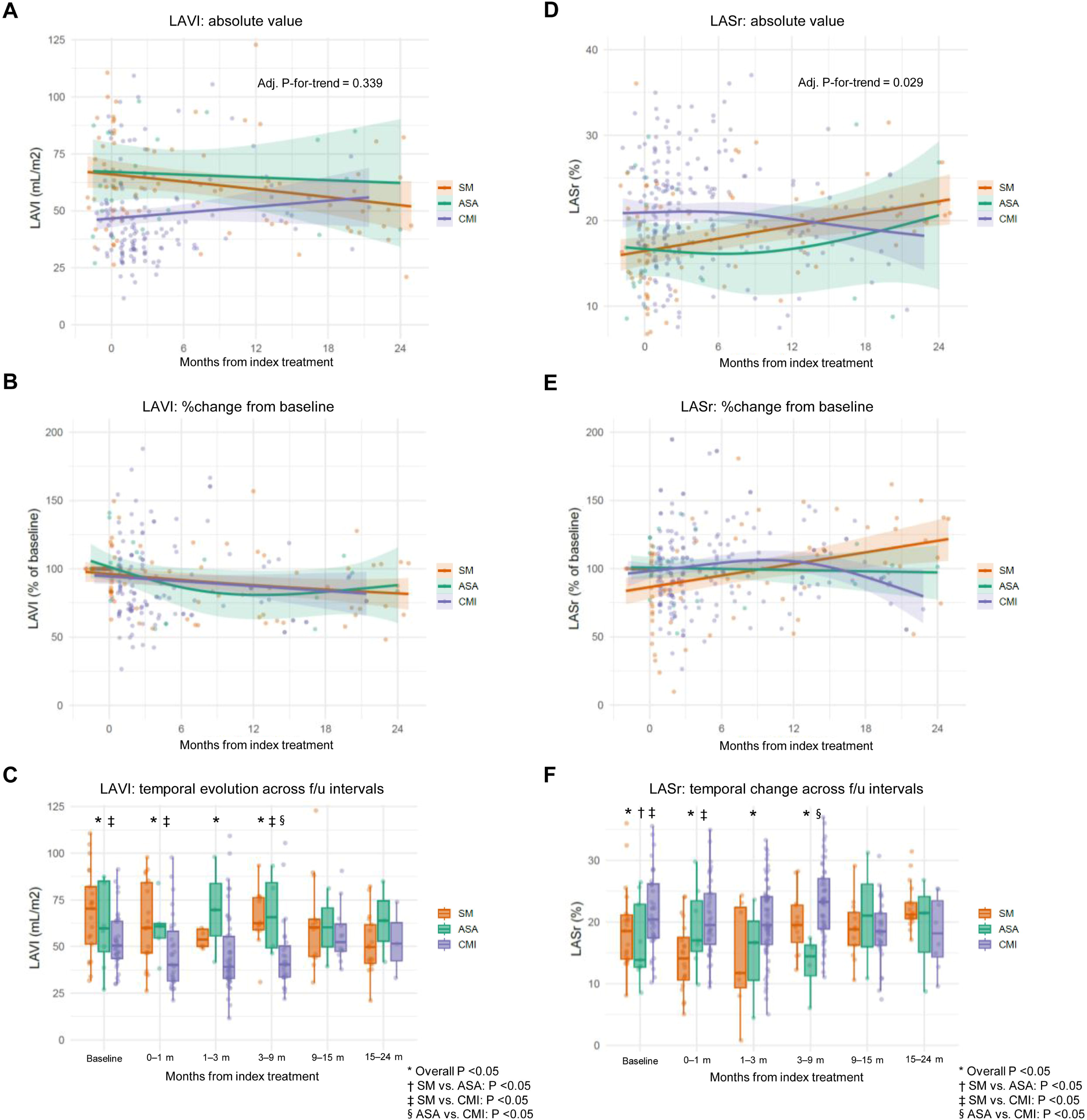
Time-trajectory of LAVI and LASr. Temporal changes in LAVI assessed as its original values (A), in specific time intervals (B), and expressed as %change from baseline (C). Temporal changes in LASr as its original values (D), in specific time intervals (E), and expressed as %change from baseline (F). * Overall P values were calculated with generalized additive mixed models (GAMM), comparing the trajectory of echocardiographic variables with adjustment for age, sex, comorbidities at baseline, and the baseline echocardiographic parameters.

In contrast, LASr demonstrated significant differences in longitudinal patterns among treatment groups (adjusted P-for-interaction = 0.029; **Figures 4D–4F**). In the SM group, LASr improved progressively over time, both as absolute values and percentage change from baseline. The ASA group showed minimal interval change in LASr. In the CMI group, LASr remained stable during the first 9 months but exhibited a subtle and gradual decline during longer-term follow-up beyond 9–12 months.

### Sensitivity analysis in total study population

Sensitivity analyses including all treated patients, irrespective of treatment success, demonstrated consistent patterns in the trajectories of LVOT gradient, LVEF, LVGLS, LAVI, and LASr (**Supplementary Figures S1–S3**).

## Discussion

In this study, we compared longitudinal LV and LA deformation after SM, ASA, and CMI therapy in patients with oHCM. Although LVOT gradient reduction was comparable across strategies, myocardial adaptation differed. SM and ASA were associated with progressive improvement in LVGLS, whereas CMI therapy showed a subtle early decline in LVEF and stable LVGLS over 24 months. LASr improved after SM but demonstrated late attenuation beyond 9–12 months in the CMI group. As the first study directly comparing LVGLS and LASr trajectories across these three treatment modalities, our findings highlight treatment-specific remodeling patterns and underscore the importance of serial deformation imaging. Notably, although CMI achieved the highest rate of hemodynamic success, improvement in deformation indices was more consistent after conventional SRT. These observations suggest that, in selected patients requiring durable long-term treatment, mechanical relief of obstruction may provide more sustained improvement in myocardial function, particularly when procedural success is anticipated to be high.

### Temporal changes in LV function after SRT or CMI

Previous studies have demonstrated that relief of LVOT obstruction by SM leads to symptomatic improvement, reductions in LVOT gradient, regression of LA volume and LV mass.(10, 21) However, LVGLS remained unchanged at mid-term follow-up, reflecting opposing regional effects—reduced longitudinal strain at the myectomy site due to resection of contractile myocardium and improved strain in non-septal segments secondary to unloading.(8, 10, 22)

After ASA, early septal strain reduction reflects localized necrosis, whereas global systolic indices are relatively preserved.(23) Indeed, the magnitude of early septal strain impairment predicts sustained gradient reduction.(7) Although long-term studies demonstrate persistent gradient reduction and LV mass regression, LVGLS frequently fails to normalize, reinforcing the concept that oHCM represents intrinsic myocardial disease rather than pure afterload excess.(6) Layer-specific strain analyses further refined these observations, demonstrating selective improvement in endocardial strain within the basal septum, with persistent impairment in mid-myocardial and epicardial layers.(24)

Importantly, prior studies have largely assessed LV function immediately or within 1 year after SRT.(8–10, 21–23) In contrast, patients receiving CMI undergo structured serial imaging, allowing more detailed temporal assessment. Substudies from VALOR-HCM and other cohorts reported modest improvement in LVGLS up to 56 weeks of mavacamten therapy.(3, 4, 25) However, heterogeneous imaging schedules across treatment modalities have limited direct comparison of deformation trajectories.

Building on prior literature, our direct comparison of temporal echocardiographic trajectories over 2 years demonstrated gradual improvement in LVGLS after SM or ASA, whereas LVGLS remained unchanged with CMI despite effective gradient reduction. Although LVEF decreased slightly across all groups, the relative decline was more pronounced with CMI when expressed as percent change from baseline. These findings suggest that mechanical unloading after SRT may facilitate recovery of global longitudinal function, whereas pharmacologic sarcomere inhibition may attenuate this adaptive response.

### Effects of SRT and CMI on LA structure and function

Data on LA remodeling after SRT is inconsistent. Some studies report reductions in LA volume with unchanged LASr,(11) whereas others demonstrate early improvement in both parameters.(26) Early prospective data following mavacamten showed reductions in LVGLS and LASr at 30 days, potentially reflecting direct negative inotropic effects on atrial myocardium.(27) Longer-term studies suggest progressive LA functional recovery during sustained CMI therapy. In VALOR-HCM, LASr improved over 56 weeks, whereas LVGLS changes were modest.(3, 4) Similarly, serial analyses up to 12 months demonstrated continued LASr improvement consistent with delayed LA reverse remodeling.(25)

In contrast, our data showed reductions in LAVI and improvement in LASr predominantly after SM or ASA, but not with CMI. These findings may reflect heterogeneous atrial responses to myosin inhibition. Recent registry data indicate that LA strain improvement during mavacamten therapy depends on baseline LA stiffness, with greater benefit in patients with more advanced remodeling.(28) Thus, our observations should not be interpreted as evidence of detrimental LA effects of CMI but rather as suggesting variability in atrial adaptation. Nevertheless, given ongoing concerns regarding atrial arrhythmia and the negative inotropic mechanism of CMI, the relative attenuation of LA functional improvement observed in our cohort warrants further investigation.

### Reappraisal of conventional SRT in the era of CMI

The negative inotropic mechanism of CMI, reflected by subtle changes in LVGLS and LASr, raises considerations regarding long-term safety and durability. Approximately 5–10% of patients receiving CMI develop LV systolic deterioration or heart failure, and concerns regarding atrial arrhythmia persist.(5) These issues may be particularly relevant in younger patients who require prolonged therapy. Cost-effectiveness is another important consideration. The estimated health-benefit price benchmark for mavacamten ranges from $12,000 to $15,000 per year,(29) and the high cost of long-term therapy may reduce its economic efficiency relative to SRT under current pricing structures.(30)

However, procedural risks—including mortality and pacemaker implantation—remain important limitations of SM and ASA.(2) In our cohort, hemodynamic success (defined as post-procedural LVOT gradient <50 mmHg) was highest with CMI, followed by SM and ASA. Conversely, SM was associated with atrial fibrillation, permanent pacemaker implantation, and ventricular septal defect, while ASA demonstrated the lowest procedural success rate. These risk profiles cannot be evaluated solely from a cost-effectiveness perspective. Therefore, selection of SRT over CMI should depend on a high anticipated procedural success rate and acceptably low complication risk. Ultimately, the risk–benefit balance between conventional SRT and CMI requires reassessment with longer-term outcome data, particularly to determine the proportion of patients who may be unable to maintain CMI because of LV dysfunction or arrhythmia.

### Limitations

The present study has several limitations. First, it was a single-center retrospective analysis without a prespecified echocardiographic follow-up schedule for the SM and ASA groups. In these patients, follow-up echocardiography was performed at the discretion of the attending physician, unlike the structured imaging protocol applied in the CMI group, which may have introduced follow-up bias. Second, potential temporal bias should be considered. SM and ASA were the mainstay treatments for LVOTO before the introduction of CMI. Following reimbursement of mavacamten by the Korean National Health Insurance Service in December 2024, no additional SM or ASA procedures were performed in our institution. Third, post-procedural echocardiograms in the SM and ASA groups may have had lower image quality compared with routine follow-up studies in the CMI group. However, strain analysis was performed using a validated deep-learning–based algorithm that is largely user-independent and has been validated in HCM populations undergoing SRT or CMI therapy.(12–14)

### Conclusions

In patients with oHCM, SM and ASA were associated with gradual improvement in LVGLS and LASr, whereas CMI therapy resulted in subtle LVEF decline, stable LVGLS, and late attenuation of LASr despite effective gradient reduction. These findings emphasize the importance of serial deformation imaging after CMI and suggest that conventional SRT may provide more sustained myocardial functional improvement in selected patients with high procedural success and acceptable risk profiles.

## Acknowledgements

We respectfully acknowledge the contributions of Professor Cheong Lim, who passed away prior to the publication of this work. He made substantial contributions to the conception and design of the study, and this work is dedicated to his memory.

## Data availability statement

Data used in this study cannot be made publicly available because of the strict ethical restrictions set by the IRB of Seoul National University Bundang Hospital (https://e-irb.snubh.org). Please contact the corresponding authors or the ethics board at SNUBH for further inquiries regarding data availability within the scope permitted by the IRB.

## Author Contributions

**Conceptualization:** In-Chang Hwang, Cheong Lim, Goo-Yeong Cho, Inho Chae.

**Data curation:** In-Chang Hwang, Minjung Bak, Jiesuck Park, Hong-Mi Choi, Yeonyee E. Yoon, Goo-Yeong Cho.

**Formal analysis:** In-Chang Hwang.

**Funding acquisition:** In-Chang Hwang.

**Investigation:** In-Chang Hwang, Minjung Bak, Jiesuck Park, Hong-Mi Choi, Yeonyee E. Yoon, Sang Yoon Kim, Joon Chul Jung.

**Methodology:** In-Chang Hwang.

**Project administration:** In-Chang Hwang, Hyoung Woo Chang, Jae Hang Lee, Jun Sung Kim, Hyung Gon Je, Sang Hon Park, Cheong Lim, Goo-Yeong Cho, Inho Chae.

**Software:** In-Chang Hwang, Jiesuck Park, Yeonyee E. Yoon.

**Supervision:** In-Chang Hwang.

**Validation:** In-Chang Hwang.

**Visualization:** In-Chang Hwang, Yeonyee E. Yoon.

**Writing – original draft:** In-Chang Hwang.

**Writing – review & editing:** In-Chang Hwang, Minjung Bak, Jiesuck Park, Sang Yoon Kim, Joon Chul Jung, Hong-Mi Choi, Hyoung Woo Chang, Jae Hang Lee, Yeonyee E. Yoon, Hyung Gon Je, Jun Sung Kim, Sang Hon Park, Cheong Lim, Goo-Yeong Cho, Inho Chae, Kay-Hyun Park.

## References

1. Ommen SR, Ho CY, Asif IM, Balaji S, Burke MA, Day SM, et al. 2024 AHA/ACC/AMSSM/HRS/PACES/SCMR Guideline for the Management of Hypertrophic Cardiomyopathy: A Report of the American Heart Association/American College of Cardiology Joint Committee on Clinical Practice Guidelines. Circulation. 2024;149:e1239–e311.

2. Liebregts M, Vriesendorp PA, Ten Berg JM. Alcohol Septal Ablation for Obstructive Hypertrophic Cardiomyopathy: A Word of Endorsement. J Am Coll Cardiol. 2017;70:481–8.

3. Desai MY, Okushi Y, Gaballa A, Wang Q, Geske JB, Owens AT, et al. Serial Changes in Ventricular Strain in Symptomatic Obstructive Hypertrophic Cardiomyopathy Treated With Mavacamten: Insights From the VALOR-HCM Trial. Circ Cardiovasc Imaging. 2024;17:e017185.

4. Desai MY, Okushi Y, Wolski K, Geske JB, Owens A, Saberi S, et al. Mavacamten-Associated Temporal Changes in Left Atrial Function in Obstructive HCM: Insights From the VALOR-HCM Trial. JACC Cardiovasc Imaging. 2025;18:251–62.

5. Garcia-Pavia P, Oreziak A, Masri A, Barriales-Villa R, Abraham TP, Owens AT, et al. Long-term effect of mavacamten in obstructive hypertrophic cardiomyopathy. Eur Heart J. 2024;45:5071–83.

6. Sommer A, Poulsen SH, Mogensen J, Thuesen L, Egeblad H. Left ventricular longitudinal systolic function after alcohol septal ablation for hypertrophic obstructive cardiomyopathy: a long-term follow-up study focused on speckle tracking echocardiography. Eur J Echocardiogr. 2010;11:883–8.

7. van Ramshorst J, Mollema SA, Delgado V, van der Wall EE, Schalij MJ, Atsma DE, et al. Relation of immediate decrease in ventricular septal strain after alcohol septal ablation for obstructive hypertrophic cardiomyopathy to long-term reduction in left ventricular outflow tract pressure gradient. Am J Cardiol. 2009;103:1592–7.

8. Desai MY, Szpakowski N, Tower-Rader A, Bittel B, Fava A, Ospina S, et al. Echocardiographic Changes Following Surgical Myectomy in Severely Symptomatic Obstructive Hypertrophic Cardiomyopathy: Insights From the SPIRIT-HCM Study. J Am Heart Assoc. 2025;14:e037058.

9. Ha KE, Choi KU, Lee HJ, Gwak SY, Kim K, Cho I, et al. Effects of septal myectomy on left atrial and left ventricular function in obstructive hypertrophic cardiomyopathy. ESC Heart Fail. 2023;10:2939–47.

10. Moravsky G, Bruchal-Garbicz B, Jamorski M, Ralph-Edwards A, Gruner C, Williams L, et al. Myocardial mechanical remodeling after septal myectomy for severe obstructive hypertrophic cardiomyopathy. J Am Soc Echocardiogr. 2013;26:893–900.

11. Weissler-Snir A, Hindieh W, Moravsky G, Ralph-Edwards A, Williams L, Rakowski H, et al. Left atrial remodeling postseptal myectomy for severe obstructive hypertrophic cardiomyopathy: Analysis by two-dimensional speckle-tracking echocardiography. Echocardiography. 2019;36:276–84.

12. Park J, Yoon YE, Jang Y, Jung T, Jeon J, Lee SA, et al. Novel deep learning framework for simultaneous assessment of left ventricular mass and longitudinal strain: clinical feasibility and validation in patients with hypertrophic cardiomyopathy. J Echocardiogr. 2025;23:258–69.

13. Park J, Yoon YE, Chun EJ, Choi HM, Hwang IC, Lee HJ, et al. Endocardial versus whole-myocardial tracking global longitudinal strain analysis in patients with hypertrophic cardiomyopathy: A preliminary comparative study. PLoS One. 2023;18:e0288421.

14. Park J, Kim J, Jeon J, Yoon YE, Jang Y, Jeong H, et al. Single-View Echocardiographic Analysis for Left Ventricular Outflow Tract Obstruction Prediction in Hypertrophic Cardiomyopathy: A Deep Learning Approach. Journal of the American Society of Echocardiography. 2025;38:1115–26.

15. Authors/Task Force m, Elliott PM, Anastasakis A, Borger MA, Borggrefe M, Cecchi F, et al. 2014 ESC Guidelines on diagnosis and management of hypertrophic cardiomyopathy: the Task Force for the Diagnosis and Management of Hypertrophic Cardiomyopathy of the European Society of Cardiology (ESC). Eur Heart J. 2014;35:2733–79.

16. Lang RM, Badano LP, Mor-Avi V, Afilalo J, Armstrong A, Ernande L, et al. Recommendations for cardiac chamber quantification by echocardiography in adults: an update from the American Society of Echocardiography and the European Association of Cardiovascular Imaging. Eur Heart J Cardiovasc Imaging. 2015;16:233–70.

17. Jang Y, Choi H, Yoon YE, Jeon J, Kim H, Kim J, et al. An Artificial Intelligence-Based Automated Echocardiographic Analysis: Enhancing Efficiency and Prognostic Evaluation in Patients With Revascularized STEMI. Korean Circ J. 2024;54:743–56.

18. Park J, Jeon J, Yoon YE, Jang Y, Kim J, Jeong D, et al. Artificial intelligence-enhanced automation of left ventricular diastolic assessment: a pilot study for feasibility, diagnostic validation, and outcome prediction. Cardiovasc Diagn Ther. 2024;14:352–66.

19. Park J, Kim J, Jeon J, Yoon YE, Jang Y, Jeong H, et al. Artificial intelligence-enhanced comprehensive assessment of the aortic valve stenosis continuum in echocardiography. EBioMedicine. 2025;112:105560.

20. Hwang I-C, Kim HM, Park J, Choi H-M, Yoon YE, Cho G-Y. Machine-Learning–Derived Phenotypes of Hypertensive Patients Using Multidimensional Clinical and Echocardiographic Data Including Strain Imaging. European Heart Journal - Digital Health. 2026.

21. Tower-Rader A, Furiasse N, Puthumana JJ, Kruse J, Li Z, Andrei AC, et al. Effects of septal myectomy on left ventricular diastolic function and left atrial volume in patients with hypertrophic cardiomyopathy. Am J Cardiol. 2014;114:1568–72.

22. Wang J, Sun X, Xiao M, Zhang M, Chen H, Zhu C, et al. Regional Left Ventricular Reverse Remodeling After Myectomy in Hypertrophic Cardiomyopathy. Ann Thorac Surg. 2016;102:124–31.

23. Abraham TP, Nishimura RA, Holmes DR, Jr., Belohlavek M, Seward JB. Strain rate imaging for assessment of regional myocardial function: results from a clinical model of septal ablation. Circulation. 2002;105:1403–6.

24. Zhang J, Zhu L, Jiang X, Hu Z. Layer-specific strain analysis of left ventricular myocardium after alcohol septal ablation for hypertrophic obstructive cardiomyopathy. Medicine (Baltimore). 2018;97:e13083.

25. Hussain K, Saleh D, Tang M, Garcia C, Kislitsina O, Meng Z, et al. Effect of Mavacamten on Serial Speckle-Tracking Strain in Patients with Hypertrophic Cardiomyopathy. J Am Soc Echocardiogr. 2025;38:1234–5.

26. Lu S, Zhang J, Zhu Y, Zhou W, Cheng X, Wang H, et al. Early left atrial reverse remodelling in patients with hypertrophic obstructive cardiomyopathy receiving transapical beating-heart septal myectomy. Interdiscip Cardiovasc Thorac Surg. 2024;39.

27. Wessly P, Lazzara GE, Buergler JM, Nagueh SF. Early Observations on Effects of Mavacamten on Left Atrial Function in Obstructive Hypertrophic Cardiomyopathy Patients. JACC Cardiovasc Imaging. 2023;16:1633–4.

28. Lim J, Kwak S, Jeong MH, Cho JY, Park C, Park J, et al. The Effect of Mavacamten on Left Atrial Strain Dynamics in Obstructive Hypertrophic Cardiomyopathy. J Am Soc Echocardiogr. 2025.

29. Beinfeld M, Wasfy JH, Walton S, Sarker J, Nhan E, Rind DM, et al. Mavacamten for hypertrophic cardiomyopathy: effectiveness and value. J Manag Care Spec Pharm. 2022;28:369–75.

30. Bansal K, Chien CV, Masri A, Riello RJ, Ahmad T, Desai NR, et al. Medicare Coverage and Patient Out-of-Pocket Costs for Mavacamten. Circ Cardiovasc Qual Outcomes. 2025;18:e011331.

