## Supplementary figures and tables for "Comparison of temporal changes in left atrial and left ventricular strain after septal myectomy, alcohol septal ablation, and cardiac myosin inhibitor"

**Short title:** Time-trajectory of LA and LV strain after SRT or CMI

In-Chang Hwang, MD^a,b^, Minjung Bak, MD^a,b^, Jiesuck Park, MD^a,b^,

Sang Yoon Kim, MD^c,d^, Joon Chul Jung, MD, PhD^c,d^, Hong-Mi Choi, MD^a,b^,

Hyoung Woo Chang, MD, PhD^c,d^, Jae Hang Lee, MD, PhD^c,d^, Yeonyee E. Yoon, MD, PhD^a,b^,

Hyung Gon Je, MD, PhD^c,d^, Jun Sung Kim, MD, PhD^c,d^, Sang Hon Park, MD, PhD^c,d^,

Cheong Lim, MD, PhD^c,d*^, Goo-Yeong Cho, MD, PhD^a,b^,

In-ho Chae, MD, PhD^a,b^, Kay-Hyun Park, MD, PhD^c,d^

^a^ Department of Cardiology, Cardiovascular Center, Seoul National University Bundang Hospital, Seongnam, Gyeonggi, South Korea;

^b^ Department of Internal Medicine, Seoul National University College of Medicine, Seoul, South Korea;

^c^ Department of Thoracic and Cardiovascular Surgery, Cardiovascular Center, Seoul National University Bundang Hospital, Seongnam, Gyeonggi, South Korea;

^d^ Department of Thoracic and Cardiovascular Surgery, Seoul National University College of Medicine, Seoul, South Korea

***** Dr. Lim passed away prior to the submission of this manuscript**.**

**Supplementary Figure S1. Time-trajectory of LVOT gradient in total study population**

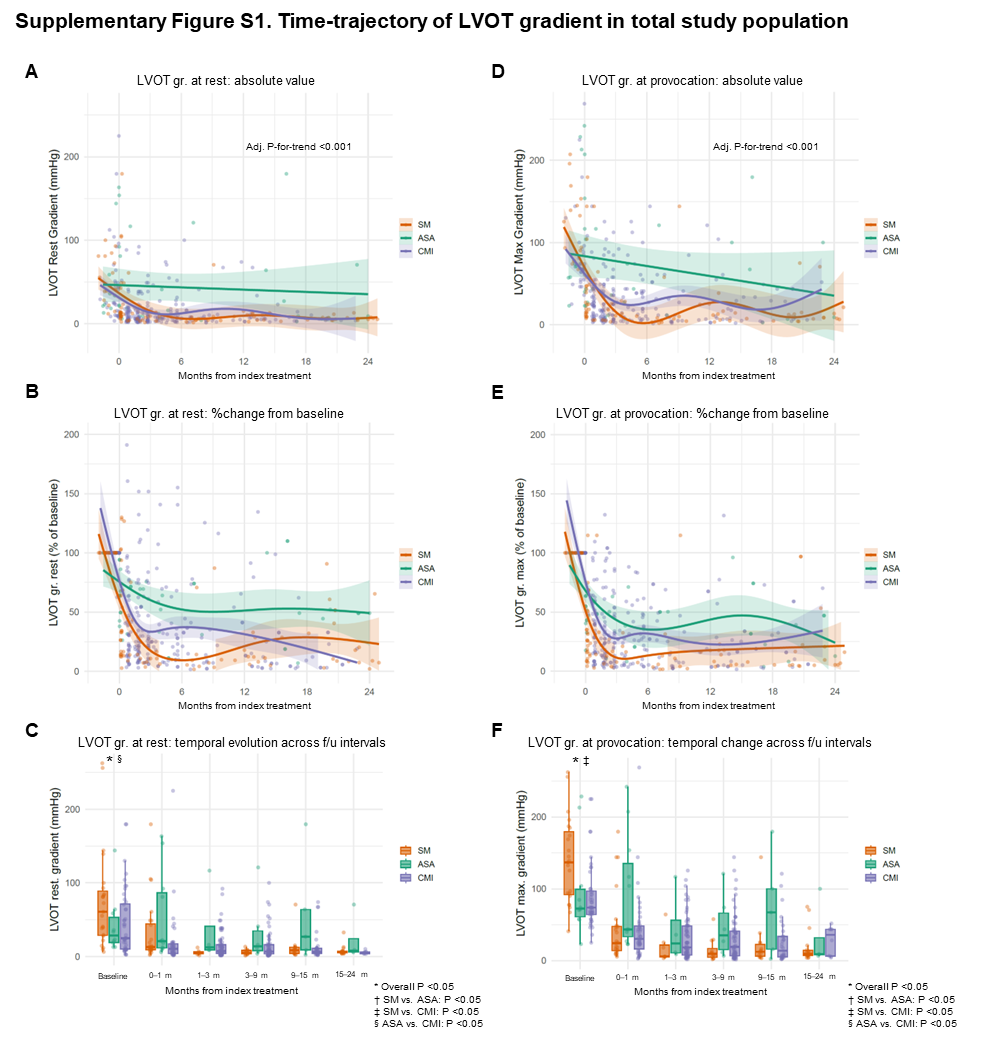

Among the total study population, temporal changes in LVOT gradient at rest assessed as its original values (A), in specific time intervals (B), and expressed as %change from baseline (C). Temporal changes in LVOT gradient after provocation as its original values (D), in specific time intervals (E), and expressed as %change from baseline (F).

* Overall P values were calculated with generalized additive mixed models (GAMM), comparing the trajectory of echocardiographic variables with adjustment for age, sex, comorbidities at baseline (hypertension, diabetes, atrial fibrillation, coronary artery disease, chronic kidney disease, and stroke), and the baseline echocardiographic parameters (LVOT gradient, LVEF, LVMI, and E/e’ ratio).

**Supplementary Figure S2. Time-trajectory of LVEF and LVGLS in total study population**

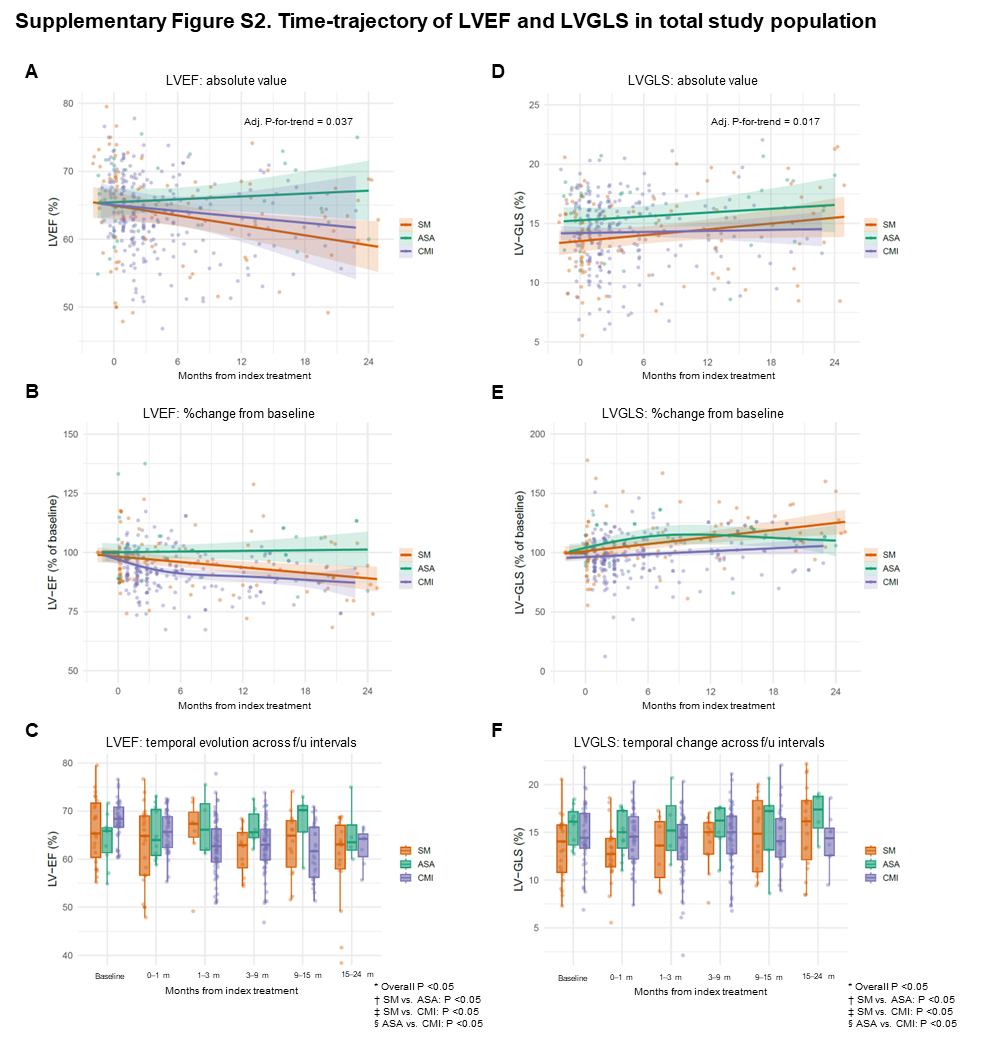

Among the total study population, temporal changes in LVEF assessed as its original values (A), in specific time intervals (B), and expressed as %change from baseline (C). Temporal changes in LVGLS as its original values (D), in specific time intervals (E), and expressed as %change from baseline (F).

* Overall P values were calculated with generalized additive mixed models (GAMM), comparing the trajectory of echocardiographic variables with adjustment for age, sex, comorbidities at baseline, and the baseline echocardiographic parameters.

**Supplementary Figure S3. Time-trajectory of LAVI and LASr in total study population**

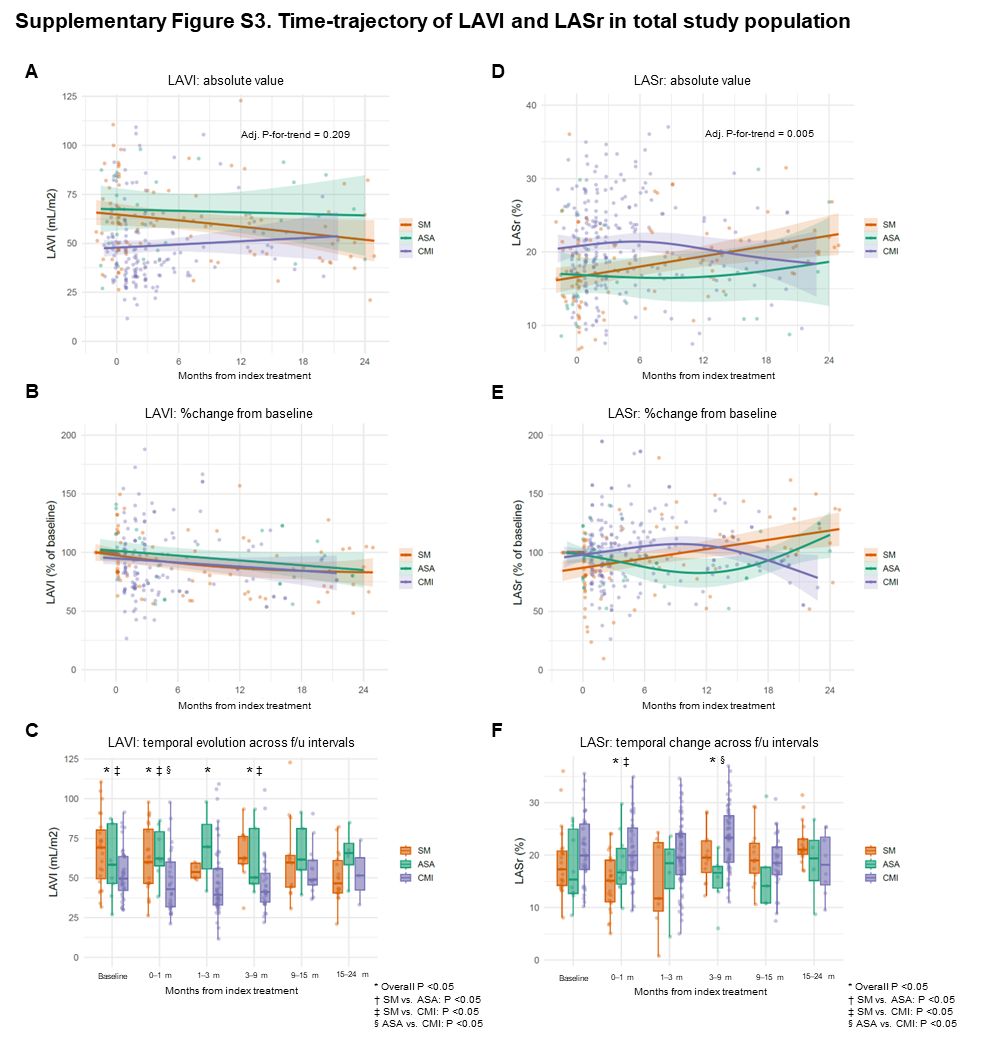

Among the total study population, temporal changes in LAVI assessed as its original values (A), in specific time intervals (B), and expressed as %change from baseline (C). Temporal changes in LASr as its original values (D), in specific time intervals (E), and expressed as %change from baseline (F).

* Overall P values were calculated with generalized additive mixed models (GAMM), comparing the trajectory of echocardiographic variables with adjustment for age, sex, comorbidities at baseline, and the baseline echocardiographic parameters.

**Supplementary Table S1. Temporal changes in echocardiographic parameters according to treatment strategies**

|  | | **Baseline** | **Post-SRT or post-CMI** | | | | |
| --- | --- | --- | --- | --- | --- | --- | --- |
|  |  |  | **<1 month** | **1–3 months** | **3–9 months** | **9–15 months** | **15–24 months** |
| **Number of patients** | **SM** | 19 | 19 | 4 | 5 | 11 | 17 |
|  | **ASA** | 8 | 8 | 2 | 4 | 2 | 3 |
|  | **CMI** | 44 | 44 | 44 | 44 | 29 | 8 |
| **LVOT gradient**  **(rest; mmHg)** | **SM** | 60.8 (29.4, 84.6) | 12.5 (9.2, 20.0) | 4.3 (3.6, 6.5) | 5.2 (3.9, 8.5) | 8.5 (4.4, 12.1) | 5.8 (5.2, 7.2) |
|  | **ASA** | 23.0 (18.3, 36.0) | 15.4 (10.6, 21.4) | 9.0 (9.0, 12.5) | 9.4 (6.1, 13.0) | 5.8 (4.1, 7.4) | 6.8 (6.3, 7.9) |
|  | **CMI** | 25.0 (10.2, 71.4) | 8.3 (4.1, 15.3) | 7.1 (4.3, 14.4) | 6.4 (3.6, 10.2) | 6.4 (4.0, 10.9) | 5.5 (4.0, 6.4) |
| **LVOT gradient**  **(provoked; mmHg)** | **SM** | 136.9 (93.3, 174.2) | 19.0 (11.8, 44.0) | 5.8 (5.6, 20.4) | 9.3 (4.8, 17.0) | 11.7 (5.7, 16.5) | 8.8 (7.1, 13.5) |
|  | **ASA** | 71.2 (57.8, 84.6) | 41.8 (26.0, 61.8) | 11.6 (10.3, 23.8) | 19.3 (10.4, 31.2) | 9.3 (5.9, 12.6) | 9.0 (7.9, 9.0) |
|  | **CMI** | 74.0 (62.4, 100.0) | 27.0 (13.7, 44.9) | 17.6 (7.0, 41.0) | 12.5 (5.8, 33.6) | 10.2 (4.8, 33.6) | 35.9 (6.5, 43.6) |
| **LVEF (%)** | **SM** | 65.4 (61.4, 72.2) | 64.4 (56.3, 69.2) | 67.4 (64.6, 69.8) | 62.9 (58.2, 65.5) | 63.8 (57.3, 68.1) | 63.4 (59.4, 67.9) |
|  | **ASA** | 65.8 (60.9, 66.7) | 62.6 (60.1, 66.0) | 62.1 (61.6, 68.8) | 64.7 (63.7, 65.4) | 64.2 (61.2, 67.2) | 62.5 (61.3, 63.5) |
|  | **CMI** | 67.8 (66.7, 70.7) | 65.9 (62.2, 69.2) | 62.2 (59.3, 66.3) | 62.5 (59.7, 66.0) | 61.2 (56.1, 66.7) | 64.2 (60.5, 65.3) |
| **LVGLS (%)** | **SM** | 15.1 (11.0, 15.8) | 12.4 (11.4, 14.4) | 13.6 (10.3, 16.1) | 15.0 (12.7, 16.0) | 13.6 (10.8, 18.6) | 16.4 (11.8, 18.4) |
|  | **ASA** | 16.4 (14.2, 17.3) | 16.3 (14.8, 17.4) | 16.8 (14.2, 18.8) | 17.5 (16.7, 17.6) | 19.2 (18.5, 20.0) | 18.7 (16.1, 18.9) |
|  | **CMI** | 15.1 (13.5, 17.1) | 14.6 (12.3, 16.5) | 14.5 (12.1, 15.8) | 14.9 (12.8, 16.7) | 14.0 (12.3, 16.3) | 14.4 (12.6, 15.4) |
| **LAVI (mL/m^2^)** | **SM** | 70.4 (51.4, 82.1) | 60.1 (46.6, 84.3) | 53.8 (50.1, 59.3) | 62.4 (58.9, 76.3) | 60.2 (44.7, 64.6) | 49.7 (41.0, 61.8) |
|  | **ASA** | 59.7 (47.2, 85.1) | 60.9 (54.5, 62.3) | 69.7 (55.8, 83.9) | 65.8 (49.3, 84.3) | 60.3 (49.9, 70.7) | 64.0 (52.9, 74.4) |
|  | **CMI** | 50.5 (43.7, 63.6) | 40.2 (31.6, 58.2) | 39.2 (33.0, 55.4) | 40.4 (33.7, 50.3) | 52.3 (47.4, 62.1) | 51.6 (42.4, 62.7) |
| **LASr (%)** | **SM** | 18.6 (14.0, 21.2) | 14.1 (10.6, 17.5) | 11.7 (9.4, 22.4) | 19.6 (16.7, 22.8) | 18.9 (16.3, 21.6) | 21.2 (20.6, 23.2) |
|  | **ASA** | 13.9 (12.7, 22.9) | 17.0 (15.3, 23.4) | 16.7 (10.6, 20.2) | 14.4 (11.3, 16.3) | 21.1 (16.0, 26.2) | 21.5 (15.1, 24.2) |
|  | **CMI** | 20.4 (17.5, 26.2) | 19.5 (16.4, 24.6) | 19.5 (16.4, 24.1) | 23.3 (18.9, 27.1) | 18.5 (16.2, 21.4) | 18.2 (14.4, 23.5) |
| **LAScd (%)** | **SM** | -10.8 (-12.5, -9.5) | -8.2 (-10.3, -6.5) | -8.1 (-12.9, -5.4) | -12.3 (-16.2, -9.2) | -11.6 (-18.1, -10.3) | -13.2 (-15.5, -12.1) |
|  | **ASA** | -9.9 (-12.2, -9.1) | -11.9 (-12.5, -11.4) | -8.9 (-10.1, -6.6) | -9.2 (-10.0, -7.7) | -15.0 (-19.4, -10.5) | -12.0 (-15.8, -10.8) |
|  | **CMI** | -13.0 (-14.5, -10.4) | -12.8 (-15.7, -9.9) | -12.7 (-16.2, -10.1) | -14.7 (-16.7, -11.1) | -10.1 (-13.2, -8.4) | -12.0 (-14.8, -8.6) |
| **LASct (%)** | **SM** | -6.8 (-9.0, -3.9) | -4.9 (-7.9, -2.7) | -5.2 (-6.8, -3.1) | -7.2 (-8.9, -4.8) | -5.1 (-6.9, -4.3) | -7.6 (-10.6, -6.2) |
|  | **ASA** | -4.1 (-7.3, -2.4) | -5.1 (-10.3, -4.1) | -7.7 (-10.1, -4.0) | -4.0 (-5.4, -3.3) | -6.1 (-6.7, -5.5) | -7.1 (-8.3, -3.1) |
|  | **CMI** | -8.3 (-11.8, -5.3) | -7.2 (-10.0, -5.5) | -7.0 (-9.1, -5.7) | -9.0 (-11.4, -6.2) | -8.2 (-10.5, -5.8) | -7.2 (-8.5, -5.9) |
| **LVMI (g/m^2^)** | **SM** | 143.2 (136.6, 177.4) | 120.2 (104.1, 151.0) | 130.8 (115.6, 141.9) | 152.6 (105.8, 174.3) | 146.4 (96.0, 156.9) | 117.1 (93.6, 159.0) |
|  | **ASA** | 146.1 (132.7, 161.2) | 150.3 (143.2, 169.4) | 115.8 (109.5, 121.8) | 137.6 (130.2, 151.4) | 156.5 (156.1, 156.8) | 123.1 (120.3, 126.6) |
|  | **CMI** | 127.4 (111.3, 142.9) | 122.6 (114.9, 140.3) | 126.0 (111.4, 143.3) | 114.0 (98.2, 125.7) | 135.2 (112.2, 165.4) | 111.8 (97.2, 126.6) |
| **E/e’ ratio** | **SM** | 19.0 (16.5, 21.8) | 18.8 (14.2, 25.3) | 32.1 (18.8, 36.8) | 18.5 (16.5, 27.0) | 21.9 (12.8, 30.7) | 15.3 (13.1, 28.4) |
|  | **ASA** | 19.9 (13.1, 32.3) | 15.2 (10.2, 24.3) | 18.1 (16.4, 29.9) | 35.8 (24.3, 47.3) | 35.8 (24.3, 47.3) | 14.9 (13.4, 19.9) |
|  | **CMI** | 18.1 (14.0, 26.4) | 14.1 (12.3, 21.3) | 15.7 (10.7, 21.4) | 14.2 (10.0, 19.0) | 24.0 (15.8, 32.5) | 19.7 (8.3, 23.8) |

Temporal changes in echocardiographic parameters are shown in patients in whom the three treatment groups (septal myectomy [SM], alcohol septal ablation [ASA], and cardiac myosin inhibitor [CMI]) were successful (defined as the maximum LVOT pressure gradient <50 mmHg within 6 months after SM, ASA, or CMI). Serial measurements were categorized across specific time intervals for descriptive visualization: baseline, post-SRT or post-CMI <1 month, 1 to 3 months, 3 to 6 months, 6 to 12 months, 12 to 18 months, and 18 to 24 months.

Values are shown in median with interquartile ranges (Q1, Q3).

Abbreviations: SM, surgical myectomy; ASA, alcohol septal ablation; CMI, cardiac myosin inhibitor; SRT, septal reduction therapy; LVOT, left ventricular outflow tract; LVMI, left ventricular mass index; LVGLS, left ventricular global longitudinal strain; LASr, left atrial reservoir strain; LAScd, left atrial conduit strain; LASct, left atrial contractile strain; LVEF, left ventricular ejection fraction; E/e′, ratio of early mitral inflow velocity to early diastolic mitral annular velocity.
